# Exploring the opportunities for improving the accessibility of rehabilitation services in rural communities in Uganda through the inclusion of CBR graduates at the primary health care level

**DOI:** 10.1101/2025.02.27.25322759

**Authors:** F Beckerlegge, S Asthana

## Abstract

**Introduction:** The increasing burden of disabilities and the need for rehabilitation services are significantly impacted by the ageing population, the rise of non-communicable diseases, and global conflicts, particularly in low and middle-income countries. The limited number of trained Allied Health Professionals for rehabilitation (Physical therapist, Occupational therapist, Speech therapist) in sub-Saharan Africa has prompted the rise of Community-Based Rehabilitation programmes to bridge this gap, particularly in rural and hard-to-reach communities.

**Aim:** To explore opportunities to improve access to clinical rehabilitation services in rural Ugandan communities by including Community-Based Rehabilitation (CBR) graduates in primary healthcare teams.

**Method:** This research used a case study approach, engaging 15 CBR graduates, Allied Health Professionals (AHPs), and CBR programme implementers in semi-structured interviews to understand the perceptions and experiences of CBR graduates’ delivery of basic rehabilitation services and the incorporation of CBR services into primary health settings to increase access to rehabilitation services in rural and hard-to-reach Ugandan communities. Thematic Analysis was used to examine and interpret the data.

**Results:** The research revealed several key issues. 1) role confusion among stakeholders about how CBR graduates compare to other healthcare roles. 2) training discrepancies with notable variations across academic institutions and different organisations, 3) lack of globally recognised roles, responsibilities and competencies for CBR graduates hinders professional recognition, and 4) concerns regarding inadequate practical skills training and heavy theoretical curriculum.

**Conclusions and limitations:** There is an urgent need for professionals to provide holistic health and social care in resource-constrained contexts, including disability awareness, empowerment, community participation, social inclusion, rehabilitation, and health education. With curriculum enhancement and additional practical skills training, Ugandan CBR graduates may be able to provide these services under the supervision of rehabilitation therapists. A fundamental shift in government priorities and funding is also needed to create opportunities and address the shortages of human resources for health and rehabilitation services.

While research on CBR programmes is scarce, this study delves into important questions related to their effectiveness. We employed diverse recruitment strategies and piloted the interviews to minimise bias resulting from the small sample size of the study. These proactive measures enhance the validity of our findings, providing a solid foundation for further study and insights into CBR programmes.

## Introduction

The ageing population, rise in non-communicable diseases, and global conflicts contribute to increasing impairments and declines in physical function. The 2019 Global Burden of Disease Report states that 1.3 billion people worldwide have significant disabilities, and 2.4 billion could benefit from rehabilitation services (WHO, 2022), marking a 63% global increase over 30 years (Cieza et al., 2020). Low and middle-income countries bear 80% of the disability burden, with one in five Africans needing rehabilitation services and years lived with disability (YLDs) increasing by 125% over 20 years to 27 million (IHME, 2023).

The World Health Organization (WHO) defines disability as the “*interaction between individuals with health conditions (e.g., cerebral palsy, Down syndrome, low back pain, and depression) and personal/environmental factors (e.g., negative attitudes, inaccessible transportation/buildings, and limited social support)”* (WHO, 2023). The link between poverty and disability is well-established, with those in poverty more likely to have a disability and vice versa (MGLSD, 2020). The intersection of poverty, disability and rurality increases disability prevalence and creates barriers to health, education, and economic engagement (Ned *et al., 2020;* Mutwali & Ross, 2019).

Rehabilitation involves interventions to improve function and reduce disability in individuals with health conditions (WHO, 2023). It includes physical, mental, and social support, essential for Universal Health Coverage. Accessible, affordable rehabilitation is crucial for maintaining independence and enhancing quality of life (Heinemann, 2020). Allied Health Professionals (AHPs) deliver rehabilitation services such as physical therapy (PT), occupational therapy (OT), speech therapy (SLT), dietetics, orthopaedics, and assistive technology (AT). However, in Uganda, there is a shortage of AHPs and other rehabilitation therapists.

Uganda, a low-income East African nation with a population of 45.6 million (World Bank 2023), faces a growing need for healthcare and rehabilitation services. Disability estimates vary greatly by source, from 8.4% (MGLSD, 2020) to 12.4% (UBOS, 2016). Stigma and cultural misconceptions often deny people with disabilities their rights to health, education, and leisure. Uganda ranks 173 out of 188 countries in the WHO health matrix for SDG 3 Universal Health Care progress (MoH, 2020). The 2022 STARS report (MoH, 2022) concluded Uganda’s rehabilitation workforce as underdeveloped and under-resourced, highlighting the urgent need for training, employment, fair deployment, retention of rehabilitation professionals, and infrastructural development to address these issues. It revealed minimal national funding priorities for rehabilitation and AT, leading to high reliance on out-of-pocket payments and donor funding, particularly for maternal and child health care. Government-supported rehabilitation services are primarily available at national and regional referral hospitals, while Faith-based private facilities and non-governmental organisation (NGO) facilities reach a few rural and hard-to-reach communities. Cuts in international financing halted government-supported CBR programmes nationally, leaving rural individuals with disabilities without essential services (MGLSD, 2020).

In 2011, only 18.5% of Ugandan children with disabilities had access to rehabilitation therapists in their communities (ACPF 2011). By 2017, this had dropped to just 10% receiving rehabilitation services, with only 3% using assistive devices (MGLSD, 2020). As of 2016, Uganda had 216 registered physiotherapists, 75% of which were in major urban areas (Nanyunja F. *et al.,* 2017), resulting in less than 1 physiotherapist per 180,000 people. In 2022, the Allied Health Professions Council (AHPC) registered only 1,022 rehabilitation professionals, mainly in secondary and tertiary care, including 381 physiotherapists, 207 occupational therapists, 383 orthopaedic technologists, and 22 speech and language therapists (MoH, 2022).

For over 40 years, the WHO has recommended community-based rehabilitation (CBR) to improve social and medical rehabilitation for people with disabilities in low-resource settings (WHO, 2023). CBR programmes use local resources and community participation to deliver cost-effective, sustainable solutions, especially in rural areas (WHO, 2023). The WHO’s CBR matrix (2010) guides CBR project execution and evaluation through five pillars: Health, Education, Livelihoods, Social, and Empowerment (see Figure 1). This holistic approach focuses on the needs and strengths of individuals with disabilities and their families while promoting social inclusion and equality (WHO 2010).

**Figure 1:**
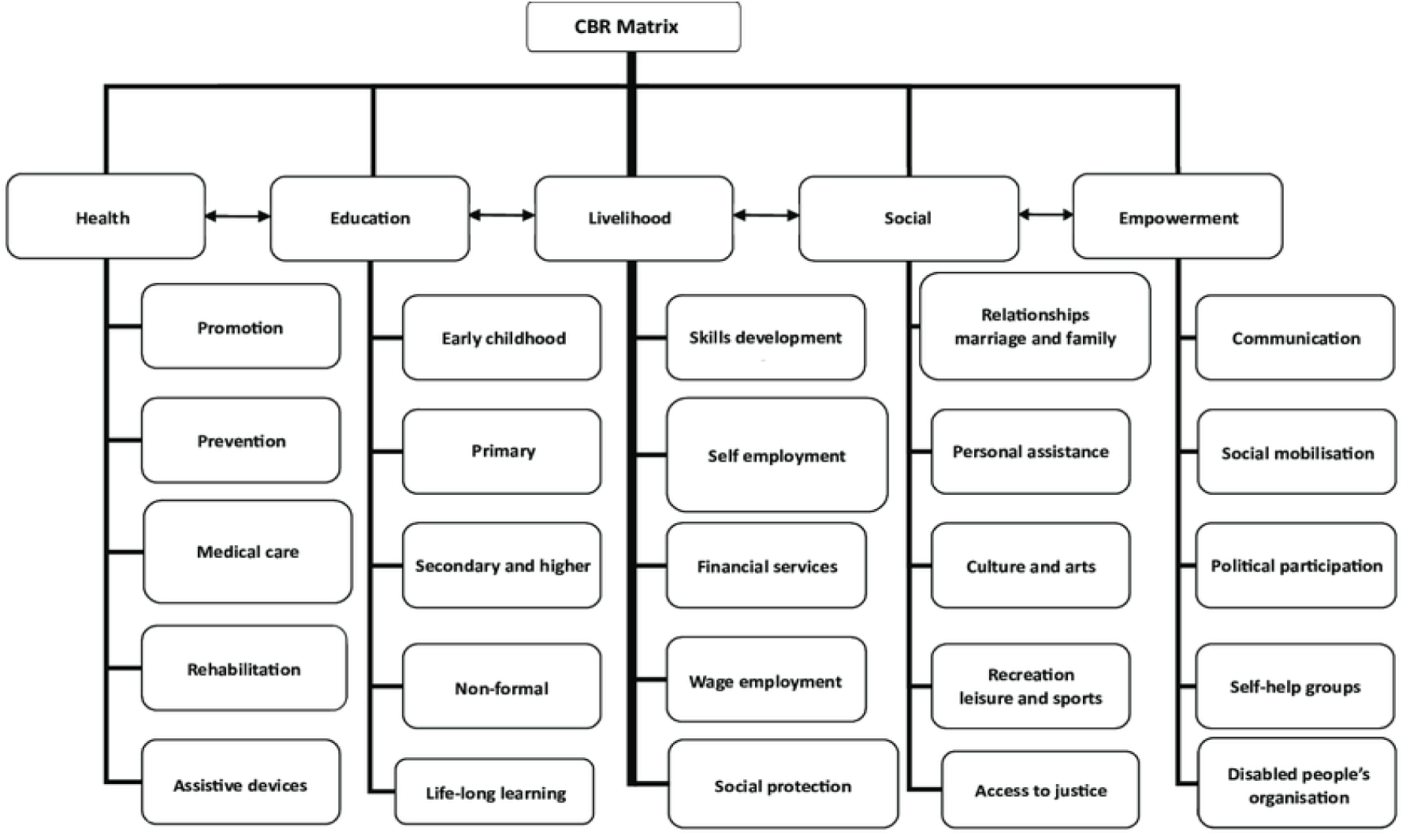
The CBR Matrix (WHO, 2010)

Uganda has attempted to address this shortage through programs like the three-year BSc in Community-Based Rehabilitation at Kyambogo University and an 18-week course offered by a non-profit Community-Based Rehabilitation Alliance (COMBRA). Both programmes provide a blend of rehabilitation and community development courses aimed at empowering graduates to work with persons with disabilities and other vulnerable individuals and groups in communities, training students to examine economic, social, and political forces marginalising vulnerable populations and with a brief introduction to physiotherapy, occupational therapy, speech therapy, and sign language (Kyambogo, 2023; COMBRA, 2022). However, neither course teaches basic clinical rehabilitation skills, and there is no clear career path or professional admission into health or social systems for these graduates. There are currently no evaluations of the course content, skill competency or impact by graduates of either course. A broad-skilled generalist cadre for CBR is seen as a way to provide low-income countries with human resources to increase access to rehabilitation services by supporting AHPs in programme delivery in rural communities (WHO, 2011).

The global lack of skilled and accessible healthcare workers threatens population health and equity (Orkin et al., 2021). Mannan *et al*. (2012) and Van Vuuren and Aldersey (2019) noted that CBR initiatives demand additional personnel with broader distribution and new skills due to the global scarcity of competent rehabilitation practitioners. Comprehensive data on CBR implementation challenges and effectiveness is lacking (Ayalew *et al*., 2020). However, evidence shows that Community Health Workers (CHW) strengthen health intervention uptake and outcomes, including immunisation uptake, childhood morbidity and mortality reduction, breastfeeding promotion, and tuberculosis treatment outcomes (Lehmann *et al*., 2007; Lewin *et al*., 2006); improve the lives of Persons with Disabilities (PWDs) (Lemmi *et al*., 2015; WHO, 2013); increases access to services, promotes social inclusion, and empowers PWDs to participate in community decision-making (Lemmi *et al*., 2016). In the African and Asian context, successful CBR implementation in Southern Africa, Cameroon, and Vietnam addressed healthcare personnel shortages and improved access to rehabilitative treatments in remote areas. However, training programmes vary across countries, with no standardisation of skills or competencies (Ned *et al*., 2020; Rule 2013).

## Aim and Objectives

The overwhelming evidence for the need to provide rehabilitation services at all healthcare levels to meet the universal healthcare targets prompts this study. This study aims to evaluate the prospective inclusion of CBR graduates in primary healthcare to support the work of allied health professionals in Uganda.

### Goal

The overall goal is to improve the accessibility of rehabilitation services in rural Uganda.

#### Research Question

What are the opportunities for incorporating CBR graduates to support the work of allied health professionals at the primary health care level to increase access to rehabilitation services in rural Ugandan communities?

#### Objectives

To explore the opportunities for improving the accessibility of rehabilitation services in rural communities in Uganda through the inclusion of CRB graduates to support the work of allied health professionals at the primary health care level.

1. To study the perceptions of allied health professionals, CBR graduates, and programme implementers regarding the feasibility and challenges of incorporating the CBR graduates to support the work of allied health professionals at the primary health care level.
2. To study the perceptions of allied health professionals, CBR graduates, and programme implementers regarding the potential impact of including CBR graduates to support the work of allied health professionals at the primary health care level.

## Methodology

### Study Design

This study uses a qualitative study design and a case study approach (Yin, 2009) for an in-depth, multi-faceted exploration of the complex issues of CBR integration in primary health settings. It examines CBR implementation across varied contexts and allows for potentially more robust and generalisable findings. The research adopts a critical epistemology, which examines existing structures and barriers that affect rehabilitation service delivery. Through this lens, the study investigates the perceptions and experiences of CBR graduates and AHPs about CBR practices and the potential for CBR inclusion into primary health care in Uganda.

### Data Collection

*Literature review and key informant interviews (KIIs) constitute the main sources of data collection for this study.* We started the study with a comprehensive review of existing literature on CBR in sub-Saharan Africa using relevant scholarly articles, books, reports, and other sources. Inclusion and exclusion criteria were applied to filter and select literature matching the research objectives (see Figure 2). Full-text research articles from 2008 to 2023, available in English and focused on sub-Saharan Africa and CBR programmes or training, were sourced from PubMed, Google Scholar, and MEDLINE, along with literature from reference lists and websites like WHO and World Bank, National Ministries, and NGOs. The literature search used keywords with Boolean and MeSH terms: community-based rehabilitation, community rehabilitation worker, community rehabilitation facilitator, low-income country, low-resource setting, Uganda, South Africa, Southern Africa, disability, human resources for health, and task-shifting. Relevant data were reviewed and extracted, identifying patterns, themes, and gaps in existing research. (PRISMA see Annexure 1).

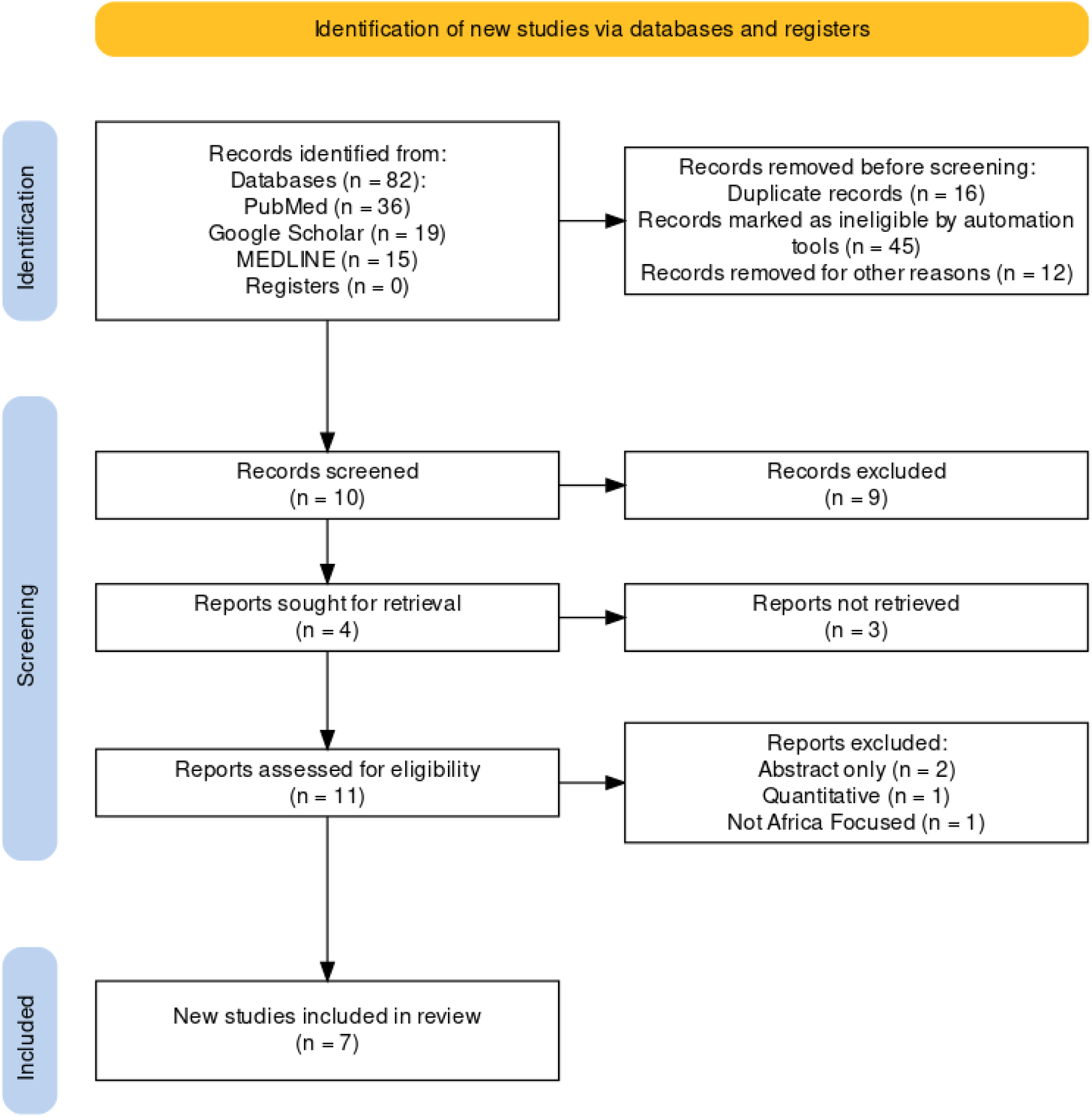

**Figure 2:**
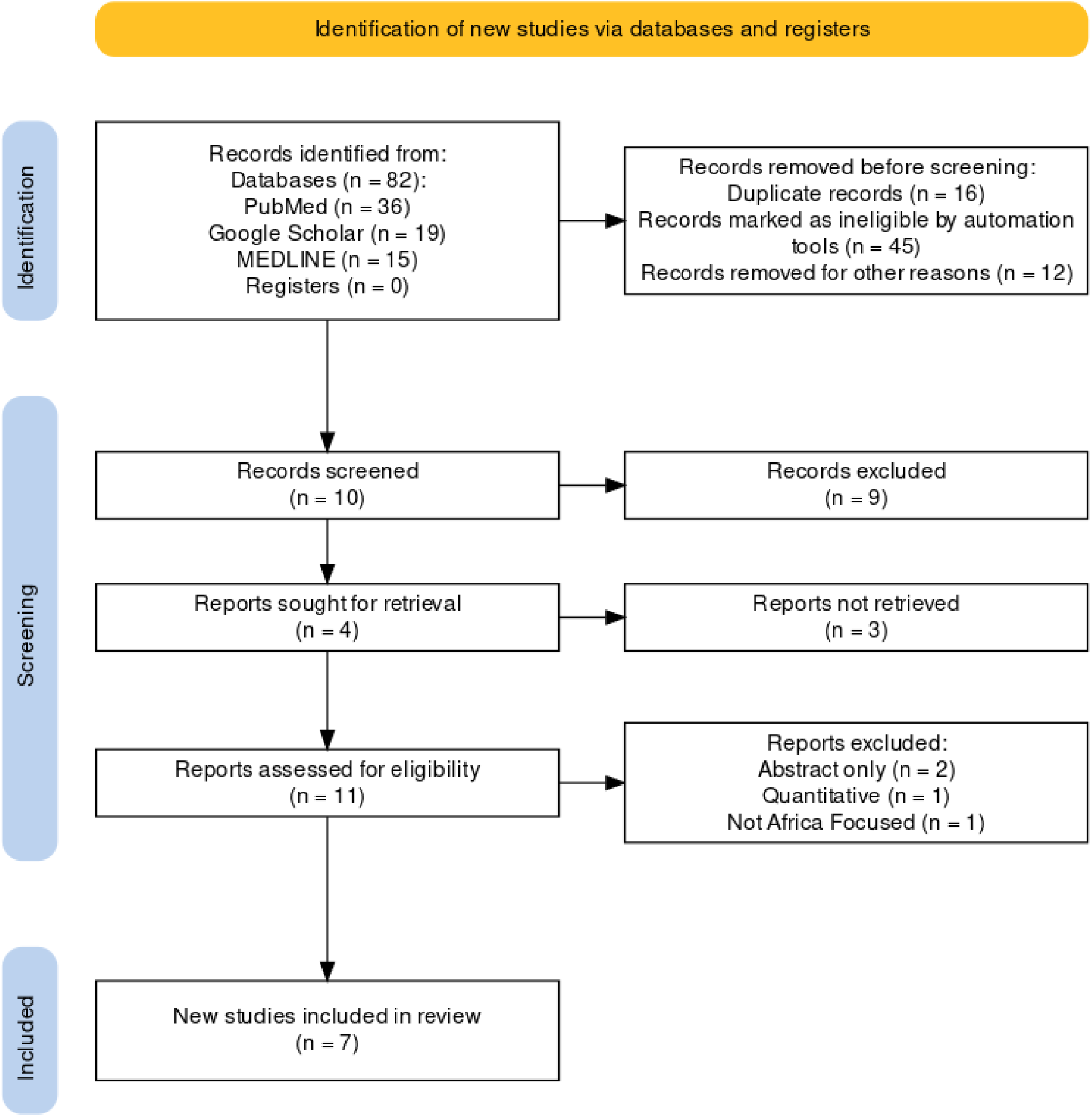
PRISMA Diagram.

Data from the review was supplemented by KIIs. A semi-structured interview guide was used to collect data from KIIs. The interview guide was designed based on the findings from the literature review, the CBR Matrix, and the researcher’s experience (Available as Annex 2). The guide ensured interviews remained focused on specific subject areas (Irvine, 2012). Interviews were conducted via Zoom or in person and audio-recorded with participant consent. Semi-structured interviews lasted between 30 and 90 minutes and were conducted between May and July 2023.

#### c) Sampling and Recruitment

25 stakeholders were purposively selected to participate in the study; out of this, 15 agreed to participate. The recruitment was based on their participation in CBR and AHP programmes, including graduation from the CBR degree at Kyambogo University, the COMBRA certificate course, or as an AHP working in community rehabilitation programmes in Uganda. Participants included one from COMBRA, four past CBR graduates from Kyambogo University, and four AHPs providing CBR services (Table 1). Through snowballing, a Ministry of Health representative in Uganda and five CBR implementers from Tanzania and South Africa were included for a comparable regional context of high poverty, limited infrastructure and minimal rehabilitation services below tertiary care level. Each participant was given an identifier code to ensure anonymity throughout the data analysis and reporting processes (Table 1).

**Table 1:** KII participants.

| Profession | Area of Work | Professional Experience | Location | ID code |
| --- | --- | --- | --- | --- |
| CBR Graduate | NGO | 8 years | Uganda | CBR01 |
| CBR Graduate | NGO | 5 years | Uganda | CBR02 |
| CBR Graduate | NGO | 2 years | Uganda | CBR03 |
| CBR Graduate | NGO | 2 years | Uganda | CBR04 |
| Speech Therapist (AHP) | NGO | 7 years | Uganda | AHP01 |
| Physiotherapist (AHP) | NGO | 4 years | Uganda | AHP02 |
| Physiotherapist (AHP) | NGO | 1 year | Uganda | AHP03 |
| Occupational Therapist (AHP) | NGO | 6 years | Uganda | AHP04 |
| CBR Implementer (AHP) | NGO | 15 years | South Africa | IMP01 |
| CBR Implementer (AHP) | NGO | 11 years | South Africa | IMP02 |
| CBR Implementer (AHP) | NGO | 2 years | Tanzania | IMP03 |
| CBR Implementer (AHP) | NGO | 8 years | Tanzania | IMP04 |
| CBR Implementer (AHP) | NGO | 5 years | Tanzania | IMP05 |
| Senior Medical Officer | Ministry of Health | 10+ years | Uganda | MOH01 |
| Trainer | COMBRA | 10+ years | Uganda | TRA01 |

### Data Analysis

All interviews were transcribed verbatim using Otter AI and checked for completeness and accuracy by the researcher. Following Braun and Clarke’s 6-step process (Nowell *et al., 2017;* Braun & Clarke, 2006), thematic analysis was used to examine and interpret data. Respondents were given unique IDs based on their affiliations to maintain anonymity.

### Ethical Considerations

Ethical approval was obtained from Queen Mary University, London, and Makerere University School of Public Health, Uganda (SPH-2023-445).

## Results

The interviews explored awareness, knowledge, and perspectives on Community-Based Rehabilitation (CBR) programmes in sub-Saharan Africa, focusing on the ability of CBR graduates to provide rehabilitation services within community health teams. The discussions yielded a variety of issues and points, summarised into four overarching themes: scope of work, skills development and training, professional considerations, and barriers to implementation. The following sections combine empirical findings with the literature review under these themes.

### Theme 1 – Scope of Work

All participants were familiar with the CBR courses and recognised the importance of holistic care for vulnerable communities. They agreed that CBR workers are well-suited for a role in the primary healthcare team to provide holistic treatment for vulnerable, hard-to-reach communities due to their training in poverty reduction, disability awareness, stigma reduction, and empowerment strategies. AHPs stated that, as specialists, they often focus more on the individual and medical perspectives, overlooking the broader issues that a CBR graduate may consider. The breadth of impact was also mirrored in the literature review.

> *”…benefits to [CBR] programme participation include physical rehabilitation, education of in-home caregivers around physical rehabilitation, emotional support and counselling, access to resources and assistive devices and increased integration into the local communities”* (Dawad & Jobson, 2011).

CBR graduates were noted for their deep understanding of local communities and cultures. AHPs recommended local recruitment of CBR graduates to a primary health team to facilitate access and engagement and support home programmes.

> *”With the concept of inclusion and CBR, I think it would be the perfect gap to fill because you are an expert in disability, but also an inclusion expert, meaning you’re leaving no one behind; those with and without disability are the vulnerable groups, even those who are not vulnerable need interventions.”* – CBR03.

However, there was some confusion among Ugandan participants about CBR graduates’ scope of work and responsibilities, which, with similarities to social workers, caused uncertainty. The literature revealed variations in CBR worker terminology, duties, payment, and training duration in South Africa, leading to confusion among healthcare workers. Some were paid, others were not. Some had received only a few training days, while others had received one or two years. However, the descriptions of their roles and responsibilities overlapped and generally included awareness creation, community engagement and home-based therapy services. This variation confused other healthcare workers as well:

> *”When doing my first internship, I called myself CBR Doctor. Can you believe it? What does that go to show? I did not know what I was doing……. If you surveyed Kyambogo graduates right now and asked, “What are you professionally?” less than half will tell you they are rehabilitation officers, or their job is in rehabilitation. They will tell you they’re CBR workers, CBR doctors, CBR practitioners, and even the understanding of CBR workers is quite hard”* – CBR03.

Interestingly, new graduate medical and allied health professionals in South Africa provide mandatory community and rural services to strengthen the public health system and motivate people to give back, thereby replacing the CBR worker model of volunteer community health workers delivering general health and rehabilitation care. Since 2004, there has been a preference to train profession-specific therapy assistants (physio, occupational and speech therapy, etc.) rather than generic rehabilitation workers whose skills crosscut the professions and include community development. However, these methods are marked with shortcomings like a lack of focus on holistic care and continuity in care, as identified by Binken et al. in 2009: *”While these new therapy assistants are competent in implementing rehabilitation programmes, they cannot offer the holistic care that CBR encourages to empower communities to assume responsibility for their health and well-being by collectively addressing the social determinants of health.”*

> *”Rural locations are not popular choices, and you don’t have permanent therapists there, so there is no continuity of services. Every year, someone leaves, and another comes, with no overlap or proper handover……. This is why we trained our CBR workers to fill the gaps and provide ongoing care”* – IMP01.

### Theme 2 – Skills Development and Training

This theme expanded on the roles of CBR graduates in Uganda and the training they receive. CBR alums, AHPs and COMBRA trainers felt that the current BSc Course was overly theoretical, leaving graduates with insufficient practical skills to implement knowledge effectively. All participants were concerned about this lack of practical experience, while all implementers and AHPs emphasised that ongoing training was also essential. Participants recommended one year of theory and more practical application for the remaining two years. This matches van Leeuwenhoekweg’s (2019) Essential Standards study, which identified the core skills CBR workers need for practical rehabilitation activities and training families to support PWDs, while Blose et al. (2021) noted: *”Continual promotion of, as well as training and education on, CBR for healthcare professionals was imperative for the successful development and roll-out of CBR programmes in South African communities*.”

> *”They are being taught about these children [CWDs] theoretically, and some confess they have never seen a child with cerebral palsy or Downs Syndrome in that training, it’s just book work …… they see what it looks like, the symptoms, but practically, they have no experience” –* AHP01.

The data also showed that graduates with basic therapy skills could increase access to rehabilitation services under AHP supervision to reduce the burden of the health workforce crisis, particularly in low-resource settings. This widely observed lack of practical experience could be attributable to the BSc graduates’ limited fieldwork, totalling 12 weeks spread across two semesters. COMBRA alums, on the other hand, are already working in the community development sector and enrol to expand their knowledge.

One CBR graduate shared his fieldwork experience with a local government office, where he had no one to mentor him. He believes he would have gained greater experience and learning opportunities within civil society, as the local government does not have adequate structure or defined positions for CBR workers.

> ”*At university, there is a lot of theory. Our only practical sessions are sign language, braille, inclusive sports, and making assistive devices……. So, for the rest of the knowledge, I think I was blessed that I was an evening student and could get practical experience during the day [working with an NGO]*”. – CBR01

Where there is no such BSc programme, NGOs in Tanzania and South Africa have taken it upon themselves to teach CBR graduates:

> ”*We target people already working in communities. We focus on developing that knowledge and skills further, how to engage communities, and how to focus on community problems. Our students do four weeks of practical work back in the communities to apply the knowledge they have learned”* – TRA01.

These informal trainings are much shorter but focus on practical skills critical to local communities. This training includes information on medical conditions and disability types and conducting basic assessments and needs evaluations. The importance of ongoing training was stressed by all implementers and AHPs as well:

> *”That is what we have learned with our projects, especially when you work with people who don’t have a scientific or medical background. You can do theoretical training but must follow up with a mentored practical application. It really helps with the understanding”* – IMP02.

AHPs recognise the need to empower lower-level health workers in the primary health care team. They believed that they were well placed in teaching CBR graduates the necessary clinical skills, and once mastered, they would be a valuable addition to the rehabilitation team, able to reach a larger number of beneficiaries and provide additional interventions to ease therapy workloads and support families:

> *”Sometimes I see a child only once a month……I would love to see them more frequently, but it’s impossible with the area we cover and the workload. If we could have CBR workers who can visit the family twice a month and complement or supplement my visits, that would help……. I could share knowledge on what I do as we work together and follow up monthly” – AHP02.*

### Theme 3 – Professional Considerations

There were strong feelings amongst all Ugandan participants that professional regulation of CBR graduates should be under the Ministry of Health (MOH) rather than the Ministry of Gender, Labour and Social Development (MGLSD). The departments need to work more closely together to adopt a human-rights approach that crosscuts ministerial boundaries. The *MGLSD* also recommends moving leadership, coordination, implementation monitoring and evaluation to an oversight office, such as the Office of the President, to support the social and political changes rather than remain segregated in various ministries (2020).

> *”The Minister of Gender, Labour and Social Development is best placed in terms of social service provision …. But I would look at strengthening the disability desk in the Ministry of Health. And this desk would make it easier to integrate CBR and create a more holistic approach.”* – CBR03.

Most participants expressed the need for a separate professional association, mentorship, supervision, and registration for professional oversight and accountability. When asked if CBR graduates posed a threat to their professional identity, AHPs believed this was unlikely. Nonetheless, they were concerned about impersonation or liability for harm:

> *”I wouldn’t want to say that it poses a threat if we have clear guidelines. Everyone knows their boundaries…. Someone may get equipped with certain skills and wants to impersonate and assume that now they are a physiotherapist and advertises …. if there are clear guidelines, we can prevent [it]”* – AHP02.

### Theme 4 – Barriers to Policy Implementation

CBR graduates and AHPs discussed integrating CBR graduates into the health system to address health workforce shortages, particularly in rural areas. They emphasised stakeholder involvement and policy identification. Ugandan participants suggested including CBR graduates within the primary health teams at parish-level Health Centre IIIs, mentored and supervised by AHPs in District or Regional hospitals.

> *”Social service provision should go lower to the people…… It’s strongly recommended that CBR graduates be at the Health Centre III level. Unfortunately, we face cases of nurses not being available at that level. Then, the Minister of Health looks at nurses at a higher standard than CBR graduates”* – CBR03.

> *”We should have CBR graduates at least in every Health Centre III level [sic]” –* CBR04.

> *”We have six* [CBR graduates] *in total; they are divided into different villages. One can cover three or four villages”* – IMP04.

CBR graduates and AHPs deemed the current Ugandan community health structure, Village Health Teams (VHTs) are inadequate in addressing rehabilitation needs. Concerns included education and training levels and frustration with the Ministry of Health’s reliance on VHTs as a substitute for CBR, which was seen as a lack of professional recognition.

> *”The VHTs, I don’t think that we should use them much for doing therapy exercises, etc*.*, because their training, if we are comparing with a CBR graduate, is a bit low. First, I would have to invest in a lot of training. CBR graduates already have that knowledge; there is something to build on”* – AHP03.

Other implementation barriers include the government’s approach to rehabilitation services and the lack of progress in incorporating adopted policies into national and district budgets and development plans. The first Disability and Rehabilitation Strategic Plan is still in draft form but will give direction on the distribution of human resources to government-aided facilities for service delivery, including community-based rehabilitation. However, participants felt rehabilitation is not prioritised in current development plans due to poor planning, budgeting, delivery, and evaluation.

> *”When we have an official supporting document signed off by the Government of Uganda advocating that we need to have these people hired, it will be up to the availability of resources. Also, engagement with the local governments to advocate for them to hire these personnel in their various district hospitals or health centre fours”* – MOH01

> *”I think the [HRH] gap in the country is not due to few physios or rehabilitation workers who have been trained. Even the ones that are available are being underutilised because of government policies. It gives me a feeling that these people [government officials] are unaware of rehabilitation and don’t consider it important in the health sector”* – AHP02.

## Discussion

This study provides valuable insights into the integration of Community-Based Rehabilitation (CBR) graduates into primary healthcare settings in Uganda, addressing a significant gap in the literature. The findings highlight the urgent need for skilled personnel to provide comprehensive health and social care in developing communities, particularly for people with disabilities and their families.

The research reveals several key issues. The first and most important issue is the confusion of roles between CBR workers and other health workers. There is widespread ambiguity among stakeholders about the role of CBR workers and how it compares to other healthcare roles. This confusion extends to researchers, healthcare staff, programme implementers, and beneficiaries. The second is the discrepancies in training. Notable variations exist in CBR training and professional development across different organisations and educational institutions. While some organisations opt for short, practical training, academic institutions often provide longer, theory-heavy programmes with limited community engagement and clinical skills training. The third is the lack of standardisation of roles and competencies. The absence of globally recognised roles, responsibilities, and competencies for CBR workers hinders their professional recognition and integration into public health systems. Last but not least are the gaps in theory and practice. Numerous concerns exist about the inadequate translation of theoretical knowledge into practical application, echoing findings from previous studies (Gindorfer & Cornielje, 2020).

The Bachelor of Science in Community-Based Rehabilitation programme at Kyambogo University aims to equip graduates with these essential skills. However, the research uncovered significant challenges these graduates face, primarily limited job opportunities within the public and civil service sectors. Stakeholders, including alums, Allied Health Professionals, and Ministry of Health representatives, concur that while the programme emphasises theoretical knowledge and social rehabilitation, it may benefit from enhanced practical components. These findings resonate with the global context of CBR programmes in low– and middle-income countries, where inconsistencies in training and competencies persist despite efforts like the WHO’s 2010 CBR Guidelines. Previous research indicating that CBR functions vary widely across nations (Blose *et al*., 2021), with diverse titles and training durations (Gilmore *et al*., 2017). The study supports the inclusion of CBR workers in multidisciplinary teams to provide holistic rehabilitation strategies but emphasises the need for additional clinical training and ongoing supervision for Ugandan BSc graduates. The research underscores the potential of task-shifting in rehabilitation, similar to successful implementations in HIV and MCH programs (Dawad & Jobson, 2011; Chung, 2019; Mapulanga & Dlugwane, 2022). However, this requires well-structured training programmes and coordinated implementation by experienced organisations.

This study has significant policy implications. It emphasises the need to develop global guidelines and strategic planning for government rehabilitation and community-based rehabilitation (CBR) efforts. Additionally, it points to the need to develop comprehensive training syllabi, curricula, and nationally recognised standards and competencies to ensure quality in these programmes. An inclusive policy process that involves all relevant stakeholders is crucial to fostering collaboration and effectiveness. Most importantly, engaging with countries that have successfully implemented CBR programmes under national policies, such as Thailand, Ethiopia, Ghana, Zambia, and Namibia, can provide valuable insights and best practices for further development.

To address the Human Resources for Health gap in clinical rehabilitation, the study suggests two key areas for improvement: first, curriculum enhancement, which involves additional training or modifications to the course’s practical components to better align with community needs and reflect training requirements for Allied Health Professionals (PT, OT, SLT, etc.) where graduates must complete an average of 75 weeks of clinical practice as part of their training before being eligible to join the professional register; and second, policy changes that call for a fundamental shift in government priorities and funding to create more opportunities for CBR graduates and address the shortage of rehabilitation professionals.

To improve community-based rehabilitation (CBR) in Uganda, we make the following recommendations. First, enhancing the practical learning components in CBR education programmes is essential for better training outcomes. Additionally, it is crucial to train healthcare professionals in CBR principles and disability inclusion to foster a more inclusive approach in healthcare settings. Secondly, developing a national CBR policy will help integrate these programmes into the district health system effectively. Third, a registry of qualified CBR personnel should also be created to ensure regulation and accountability within the sector. Furthermore, conducting a mapping exercise of existing CBR programmes will allow for an assessment of their current impact and coverage across the country. Lastly, further studies should be undertaken to evaluate the impact and cost-effectiveness of CBR workers, using inclusive methodologies to capture diverse perspectives and outcomes.

## Conclusion

This study is the first to consider perceptions regarding the integration of Community-Based Rehabilitation (CBR) graduates into primary healthcare settings in Uganda. The findings reveal a critical and urgent need for professionals capable of delivering holistic health and social care in resource-constrained environments, with a focus on disability awareness, empowerment, community engagement, social inclusion, individualised rehabilitation, and health education.

While the study provides valuable insights, the researchers acknowledge its limitations. The scarcity of research on CBR programmes in sub-Saharan Africa, particularly outside South Africa, may impact the accuracy of the literature review. Additionally, the primary data sample size was limited, potentially restricting generalisability. To mitigate these limitations, the researchers employed diverse participant recruitment, piloted interview questions, and used thematic analysis to identify patterns while minimising social desirability bias.

In conclusion, this study highlights the critical role of CBR graduates in addressing Uganda’s rehabilitation needs and the challenges they face in professional integration. By addressing these challenges through curriculum enhancements and policy changes, Uganda can make significant strides towards improving rehabilitation services, particularly in underserved communities. This research contributes to the local understanding of CBR in Uganda and adds to the global dialogue on standardising and strengthening CBR programmes worldwide.

## Data Availability

All data produce in the present study are available upon reasonable request to the authors

## Acknowledgements

The authors are grateful to the key informants who shared their time, experiences, and expertise to conduct this research.

## Appendix 1 – PRISMA flow diagram

The review identified peer-reviewed journal articles published between 2009 and 2021 from sub-Saharan Africa. An initial 82 articles were identified, and 16 duplicate entries were removed, leaving 68 articles for review; 11 met the inclusion criteria for full-text review, which included a qualitative evaluation of CBR programmes, CBR impact evaluation, or perceptions of CBR programmes. Only 7 articles were reviewed from the initial search output of 82 articles; six were focused on CBR programmes in South Africa and one in Ethiopia. Table 1 shows the PRISMA flow diagram.

Inclusion Criteria:

- Full-text article
- Written in English or English translation available
- Geographically focused in Sub-Saharan Africa
- Published between 2008 and 2023
- Qualitative research using interviews/questionnaires
- Primary focus on the perceptions of CBR programmes and training Exclusion Criteria:
- Abstract only
- Not written in English or no translation available
- Geographically focused on the rest of the world
- Published before 2008
- Quantitative research or systematic review
- Primary focus is not on the perceptions of CBR programmes and training

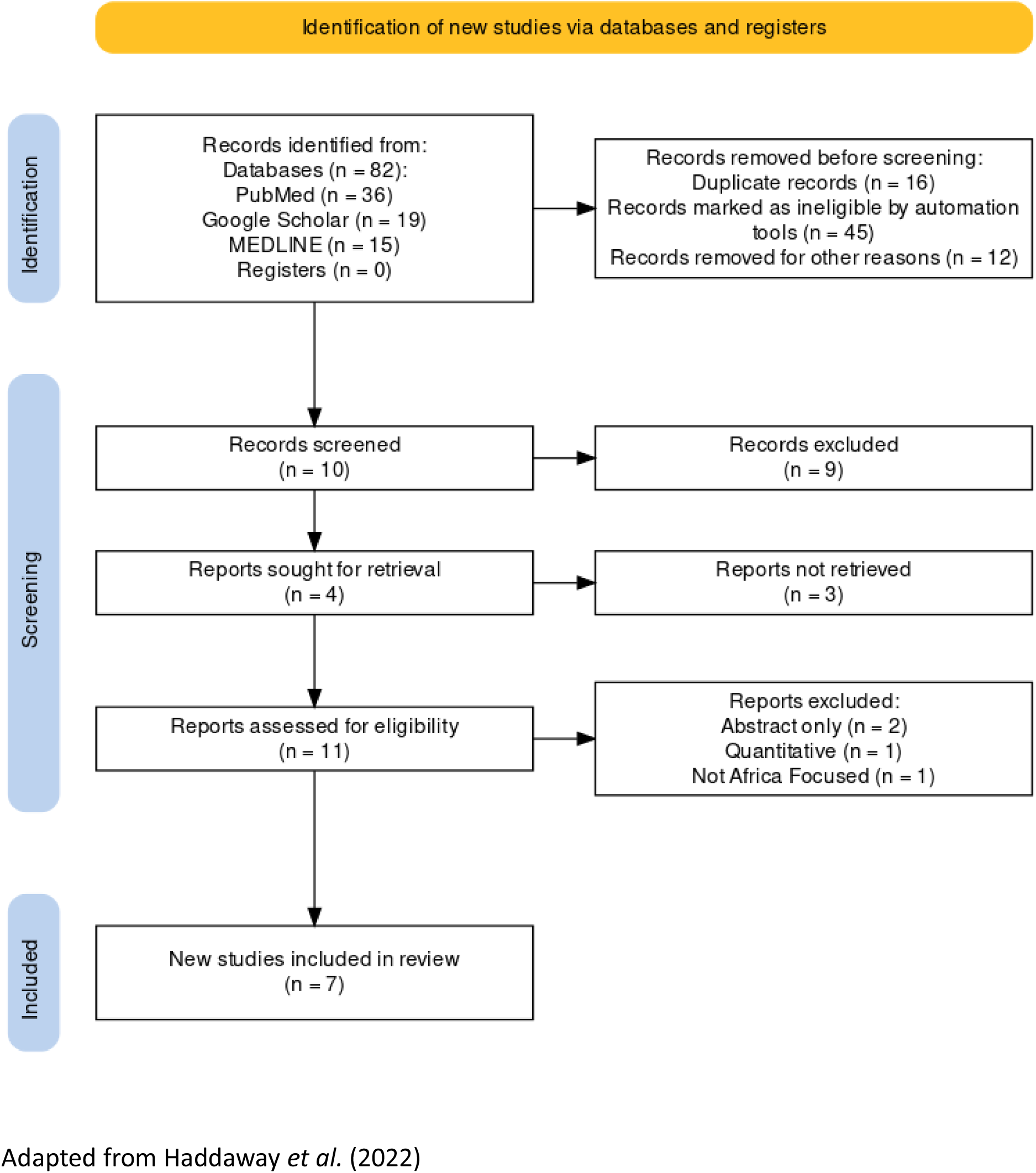

## Appendix 2 – Final full-text articles reviewed

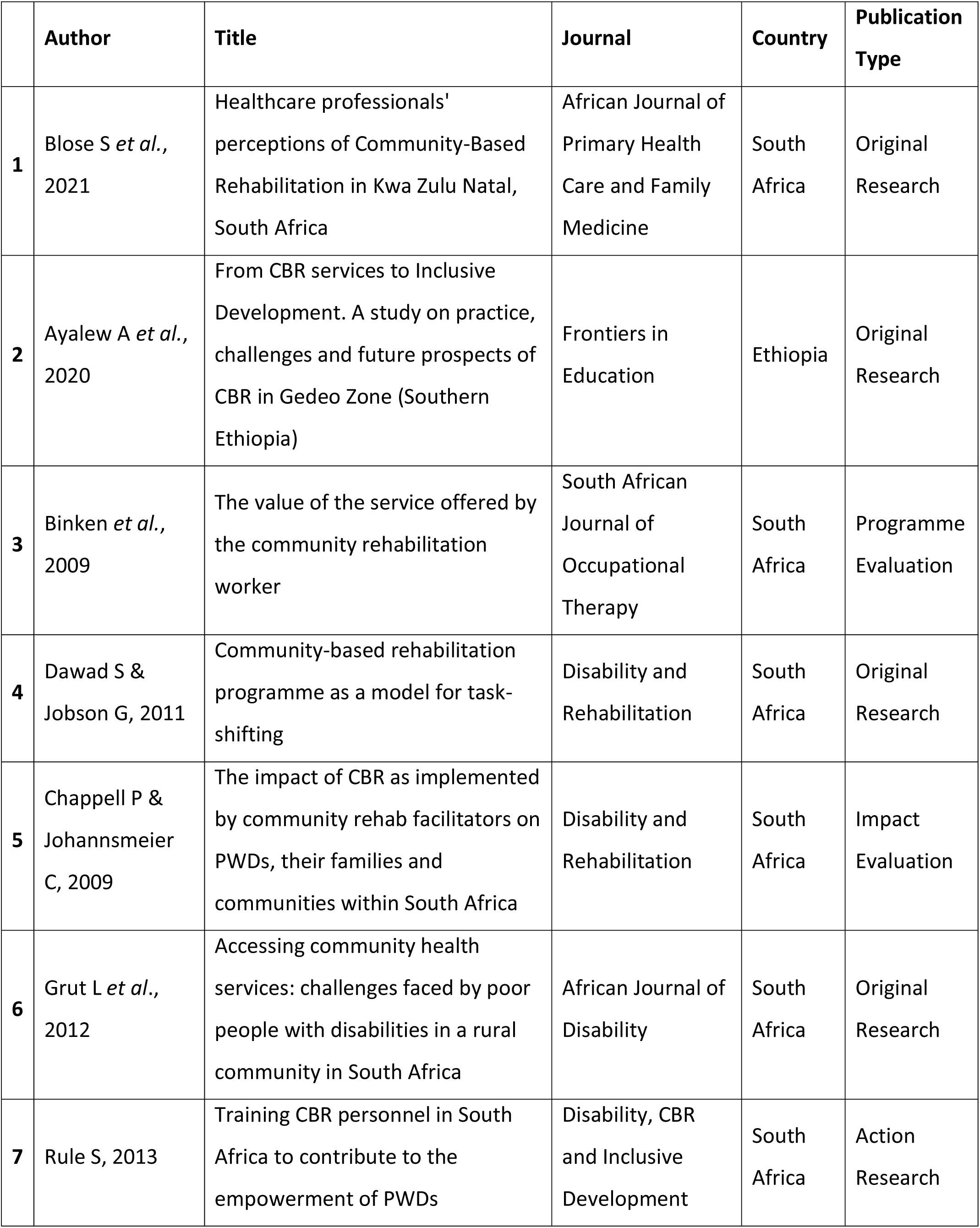

|  | **Author** | **Title** | **Journal** | **Country** | **Publication Type** |
| --- | --- | --- | --- | --- | --- |
| **1** | Blose S *et al.*, 2021 | Healthcare professionals' perceptions of Community-Based Rehabilitation in Kwa Zulu Natal, South Africa | African Journal of Primary Health Care and Family Medicine | South Africa | Original Research |
| **2** | Ayalew A *et al.*, 2020 | From CBR services to Inclusive Development. A study on practice, challenges and future prospects of CBR in Gedeo Zone (Southern Ethiopia) | Frontiers in Education | Ethiopia | Original Research |
| **3** | Binken *et al.*, 2009 | The value of the service offered by the community rehabilitation worker | South African Journal of Occupational Therapy | South Africa | Programme Evaluation |
| **4** | Dawad S & Jobson G, 2011 | Community-based rehabilitation programme as a model for task-shifting | Disability and Rehabilitation | South Africa | Original Research |
| **5** | Chappell P & Johannsmeier C, 2009 | The impact of CBR as implemented by community rehab facilitators on PWDs, their families and communities within South Africa | Disability and Rehabilitation | South Africa | Impact Evaluation |
| **6** | Grut L *et al.*, 2012 | Accessing community health services: challenges faced by poor people with disabilities in a rural community in South Africa | African Journal of Disability | South Africa | Original Research |
| **7** | Rule S, 2013 | Training CBR personnel in South Africa to contribute to the empowerment of PWDs | Disability, CBR and Inclusive Development | South Africa | Action Research |

## Appendix 3 – CASP qualitative analysis

A critical analysis of the selected literature was conducted to assess the sources’ quality, relevance, and reliability using the CASP qualitative checklist (2018). This table shows the performance of the different articles to the 10 CASP questions. None of the articles considered the relationship between the researcher and the participants (Q6), and only three could show that ethical issues had been considered (Q7). All articles were considered to be valuable (Q10).

|  |  | Clear statement of aims | Appropriate Methodology | Appropriate Design | Appropriate Recruitment Strategy | Appropriate data collection | Participant / researcher relationship considered | Ethical issues considered | Rigorous data analysis | Clear statement of findings | Was it valuable |
| --- | --- | --- | --- | --- | --- | --- | --- | --- | --- | --- | --- |
|  | Author | 1 | 2 | 3 | 4 | 5 | 6 | 7 | 8 | 9 | 10 |
| 1 | Blose *Set al.*, 2021 | Y | Y | Y | Y | Y | N | Y | Y | Y | Y |
| 2 | Ayalew A *et al.*, 2020 | Y | Y | Y | Y | Y | C | C | Y | Y | Y |
| 3 | Binken *et al.*, 2009 | Y | Y | Y | Y | Y | C | N | Y | Y | Y |
| 4 | Dawad S & Jobson G, 2011 | N | Y | Y | Y | Y | N | C | Y | Y | Y |
| 5 | Chappell P & Johannsmeier C, 2009 | Y | Y | Y | Y | Y | C | Y | Y | Y | Y |
| 6 | Grut L *et al.*, 2012 | N | Y | Y | Y | Y | N | Y | C | Y | Y |
| 7 | Rule S, 2013 | Y | Y | C | Y | Y | N | N | Y | Y | Y |

Y – Yes N – No C – Can't tell CASP (2018)

## Appendix 4 – Interview Guide

### CBR Graduate – Interview Guide

#### Introduction

*Thank the participant for their time and briefly explain the interview’s purpose*.

*Ask for their consent to record the interview, ensuring confidentiality and anonymity.Inform them that the interview will take approximately 30-40 minutes*.

#### Background Information

Tell me about your current role and work experience.

*Encourage the participant to chat about what they do. The aim is to get them feeling relaxed*.

#### Training Program Overview

Tell me more about the CBR training program you completed. How do you think it prepared you foryour current role?

What is the role of a Community-Based Rehabilitation officer/worker in Uganda?

#### Clinical Skills Training

Tell me more about the practical and clinical skills training that you did as part of your course.

*Probing Questions: Ask about the specific clinical skills taught, such as physio, occupational and speech therapyinterventions, and sign language*.

*Explore the methods of teaching these clinical skills, such as hands-on practice, simulations, or case studies*.

*Ask about fieldwork during the course*.

#### Integration of Clinical Skills in Community Rehabilitation

Do you think CBR graduates’ role could be expanded in Uganda?

*Prompts – how, where would this be, what would this involve, would it need more resources/training*

How do you think CBR graduates could contribute to the Human Resources in Health Gap in rehabilitation?

What challenges or barriers are encountered when applying clinical skills in the community?

Ask about the participant’s perspective on the impact of clinical skills training on the overall quality of community-based rehabilitation services.

#### Professional Considerations

Do you think that CBR workers should be professionally registered? Who should hold this register?Ministry of Health of Ministry of Gender, Labour and Social Development?

What other measures should be in place to ensure effective programme implementation?

#### Closing

*Thank the participant for their valuable insights and contributions*.

*Offer the participant the opportunity to add comments or ask questions. Confirm the participant’s willingness to be contacted for further clarification.End the interview and express gratitude for their participation*.

### Allied Health Professionals – Interview Guide

#### Introduction

*Thank the participant for their time and briefly explain the interview’s purpose*.

*Ask for their consent to record the interview, ensuring confidentiality and anonymity.Inform them that the interview will take approximately 30-40 minutes*.

#### Background Information

Tell me about your current role and work experience.

*Encourage the participant to chat about what they do. The aim is to get them feeling relaxed*.

#### CBR Program Overview

Do you know anything about the CBR training programmes in Uganda (Kyambogo or COMBRA)? Haveyou ever worked with someone who has completed this course?

What is your understanding of the role of a Community-Based Rehabilitation officer/worker inUganda?

#### Clinical Skills Training

If you have worked with a graduate of the CBR course, did they have basic clinical skills that couldsupport you in your work as an OT/PT/SLT?

*Probing Questions:*

#### Integration of Clinical Skills in Community Rehabilitation

Do you think CBR graduates’ role could be expanded in Uganda?

*Prompts – how, where would this be, what would this involve, would it need more resources/training*

How do you think CBR graduates could contribute to the Human Resources in Health Gap in rehabilitation?

What benefits could arise from including CBR graduates in the rehab team?

What challenges or barriers could arise in including CBR graduates in the rehabteam?

#### Professional Considerations

Do you think including CBR workers will threaten your professional identity?

Do you think that CBR workers should be professionally registered? Who should hold this register?Ministry of Health of Ministry of Gender, Labour and Social Development?

What other measures should be in place to ensure effective programme implementation

#### Closing

*Thank the participant for their valuable insights and contributions*.

*Offer the participant the opportunity to add comments or ask questions. Confirm the participant’s willingness to be contacted for further clarification.End the interview and express gratitude for their participation*.

**Table 1:** Interview Participants.

| Profession | Area of Work | Professional Experience | Location | ID code |
| --- | --- | --- | --- | --- |
| CBR Graduate | NGO | 8 years | Uganda | CBR01 |
| CBR Graduate | NGO | 5 years | Uganda | CBR02 |
| CBR Graduate | NGO | 2 years | Uganda | CBR03 |
| CBR Graduate | NGO | 2 years | Uganda | CBR04 |
| Speech Therapist | NGO | 7 years | Uganda | AHP01 |
| Physiotherapist | NGO | 4 years | Uganda | AHP02 |
| Physiotherapist | NGO | 1 year | Uganda | AHP03 |
| Occupational Therapist | NGO | 6 years | Uganda | AHP04 |
| CBR Implementer | NGO | 15 years | South Africa | IMP01 |
| CBR Implementer | NGO | 11 years | South Africa | IMP02 |
| CBR Implementer | NGO | 2 years | Tanzania | IMP03 |
| CBR Implementer | NGO | 8 years | Tanzania | IMP04 |
| CBR Implementer | NGO | 5 years | Tanzania | IMP05 |
| Senior Medical Officer | Ministry of Health | 10+ years | Uganda | MOH01 |
| Trainer | COMBRA | 10+ years | Uganda | TRA01 |

